# Resilience following a nature-based retreat: a prospective observational study

**DOI:** 10.64898/2026.11.21.26368644

**Authors:** Raz Balin, Matan Lev, Adi Schneider Moreh, Itamar Grotto

## Abstract

**Background:** Residential nature-based programs may support resilience and well-being among people experiencing substantial psychological distress. We examined changes following an independently organized retreat.

**Methods:** This prospective observational study followed 18 adults attending a five-day resilience retreat in Finland. The study did not undertake diagnostic assessments or assign retreat participation. The primary outcome was CD-RISC-10 resilience at 21 days after the retreat. Secondary outcomes included perceived stress, affect, well-being, self-efficacy and posttraumatic stress symptoms. Analyses used mixed-effects models and paired Wilcoxon tests.

**Results:** At 21 days, the mixed model showed a 5.08-point increase in CD-RISC-10 (95% CI 2.51 to 7.65; p = 0.00011). The estimate persisted on a small-sample reference (p = 0.00034) and among the 14 participants with an intake reading (+4.59 points; p = 0.0014). Perceived stress, positive and negative affect, well-being and self-efficacy also registered improvement (each paired p < 0.01). Well-being rose by 21.66 points on the 0 to 100 WHO-5 index (95% Wald CI 14.29 to 29.03). Score improvements were largest at retreat completion, partly subsided by 14 days where measured but were sustained to the 21-day mark. Evidence for improvement in posttraumatic stress symptoms was less definitive.

**Limitations:** Uncontrolled design and a small sample limit causal interpretation.

**Conclusions:** Higher self-reported resilience and improved well-being persisted three weeks after the retreat. The findings support controlled evaluation of the program.

**Registration:** Open Science Framework, DOI 10.17605/OSF.IO/7WKM5.

## 1. Introduction

Exposure to collective trauma can have enduring effects on mental health. In a nationwide Israeli cohort assessed before and after the attacks of October 7 (2023), about one in five adults followed a vulnerable or chronic course of posttraumatic stress symptoms (Levin et al., 2025). Resilience, the capacity of one to adapt, recover and maintain sustained functioning under stress and adversity, is increasingly understood as an adjustable capacity rather than a fixed one. Scores on the Connor-Davidson Resilience Scale can increase following treatment (Connor and Davidson, 2003). Supporting resilience capacity improvement alongside symptom management is therefore a relevant and effectual public-health objective.

Several intervention paradigms address resilience capacity, among them resilience training, nature-based programs and paced-breathing practices. Meta-analyses of resilience training report pooled effects of about 0.4 to 0.5 standard deviations, across 25 randomized trials (Leppin et al., 2014), 11 randomized trials (Joyce et al., 2018) and a multilevel analysis of 268 studies, though many of them were below practical significance (Liu et al., 2020). Across 50 studies of nature-based outdoor activities, 16 randomized trials showed reduced depressive and anxiety symptoms and increased positive affect, with uncontrolled studies largely agreeing (Coventry et al., 2021). Nature-based mindfulness training showed a medium pre-post effect across 25 studies of mostly low quality (Djernis et al., 2019). Forest bathing reduced depression and anxiety across 36 studies (Siah et al., 2023). Additionally, paced breathing near six breaths per minute, the resonance frequency of the cardiovascular system (Lehrer et al., 2000), reduced stress and anxiety against control when training was combined with biofeedback instrumentation, in a meta-analysis of 24 studies (g 0.83; Goessl et al., 2017). A meta-analysis of 12 randomized breathwork trials found a smaller effect on stress (g −0.35; Fincham et al., 2023).

Residential retreat programs combine these paradigms over several days away from daily routines. A systematic review of 23 retreat studies found benefits over every retreat program but limited by small samples, poorly reported methodological rigor and scarce follow-up measurements (Naidoo et al., 2018). Improvements measured at the end of a recreational break can fade after the return to routine (de Bloom et al., 2009). Determining the enduring change is further complicated by the outcome measure. The CD-RISC is among the best-rated resilience scales (Windle et al., 2011), although a systematic review of its measurement found the evidence on its responsiveness limited (Sharif-Nia et al., 2024).

In this study, we report a prospective observational evaluation of an independently organized five-day nature-based retreat in Finland. The research assessed the existing program without appointing participation or diagnostic assessments. This resilience-focused retreat combined guided nature exposure, paced breathing training, mindfulness training, moderate physical activity and group reflection sessions, with elective sauna and cold-water immersion sessions. The primary outcome was self-reported resilience on the standardized CD-RISC-10 questionnaire at 21 days post-retreat. Secondary outcomes were perceived stress, affect, well-being, self-efficacy and posttraumatic stress symptoms; future-event specificity analysis was assessed in a subset of participants as an exploratory measure. Supplement S1 documents the departures from the pre-specified plan that apply to the analyses reported here, while section 2.5 describes the analyses. The physiological-domain OSF plan also specified a secondary analysis of the questionnaire outcomes, reported in Supplement S2; the physiological measurements are the subject of a companion paper.

## 2. Methods

### 2.1 Design and registration

This prospective observational study evaluated an existing retreat program under a Ben-Gurion University of the Negev (BGU) umbrella protocol. Participation in the retreat was not assigned by the investigators. The umbrella analysis plan designates the CD-RISC-10 as the primary outcome, assessed here at the end of the 21-day follow-up period (section 2.5). The autonomic-neuroscience plan was registered on the Open Science Framework on June 17, 2026, before data collection began (DOI 10.17605/OSF.IO/7WKM5; Amendments 1 to 6). The pre-registration governs the secondary analysis, detailed in Supplement S2. The ethics approval procedure is described in section 2.6.

### 2.2 Participants

All participants were over 18 years old. Participants self-reported significant exposure to stress. No diagnostic assessment was performed in this study. At intake, 13 of 15 PCL-5 scores were at or above the screening cutoff of 33 (Bovin et al., 2016), and 13 of 14 WHO-5 index scores were at or below 50, the index’s cutoff for poor well-being (Topp et al., 2015). Pre-specified exclusion criteria were acute suicidality or a recent suicide attempt, active untreated psychosis, active severe substance-use disorder along with any condition making participation unsafe, pregnancy where thermal exposure was included, and inability to consent or to wear the measurement device (Empatica EmbracePlus bracelet). Declining research participation did not affect retreat participation. Age and sex, the registered covariates, were held on the intake record apart from the research data and were not linked to the coded dataset, to limit identification risk in a small cohort (Deviation 75). Age on the intake record averaged 35.7 years (SD 8.9; range 22 to 49).

The recruitment period was February 15 to May 17, 2026. Intake batteries were completed on July 29 and August 2, 2026, with one late completion on the first retreat day. The retreat ran from August 9 to 13, 2026. Follow-up concluded at 21 days post-retreat, on September 3. The collected dataset was frozen on September 5. Four of the 18 participants failed to complete the baseline battery. At day 21 post-retreat, 13 participants had paired CD-RISC-10 scores and 14 had paired PCL-5 scores. Table 1 and Supplement S3 document the collection cascade and instances of missing data.

**Table 1.**
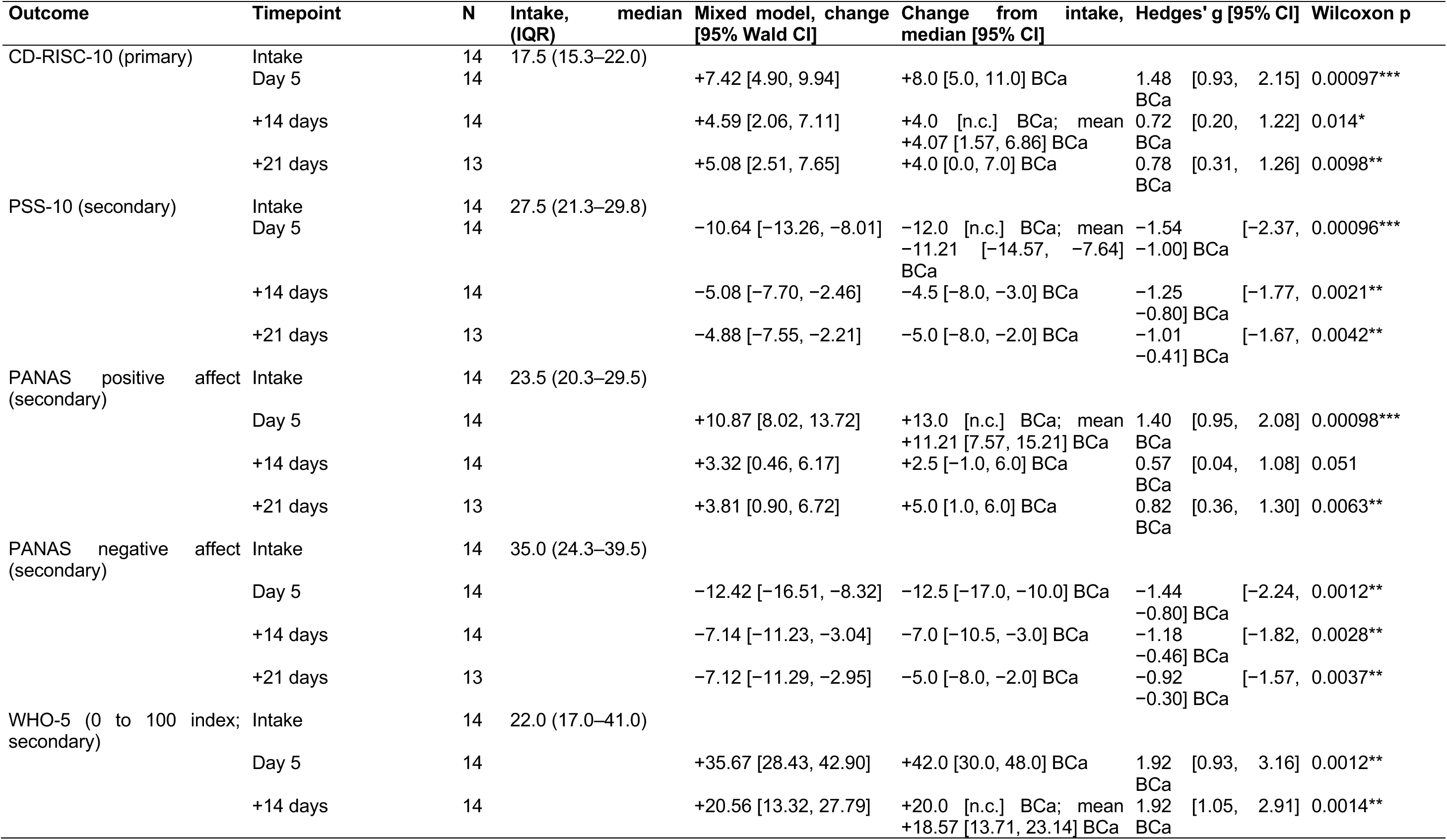
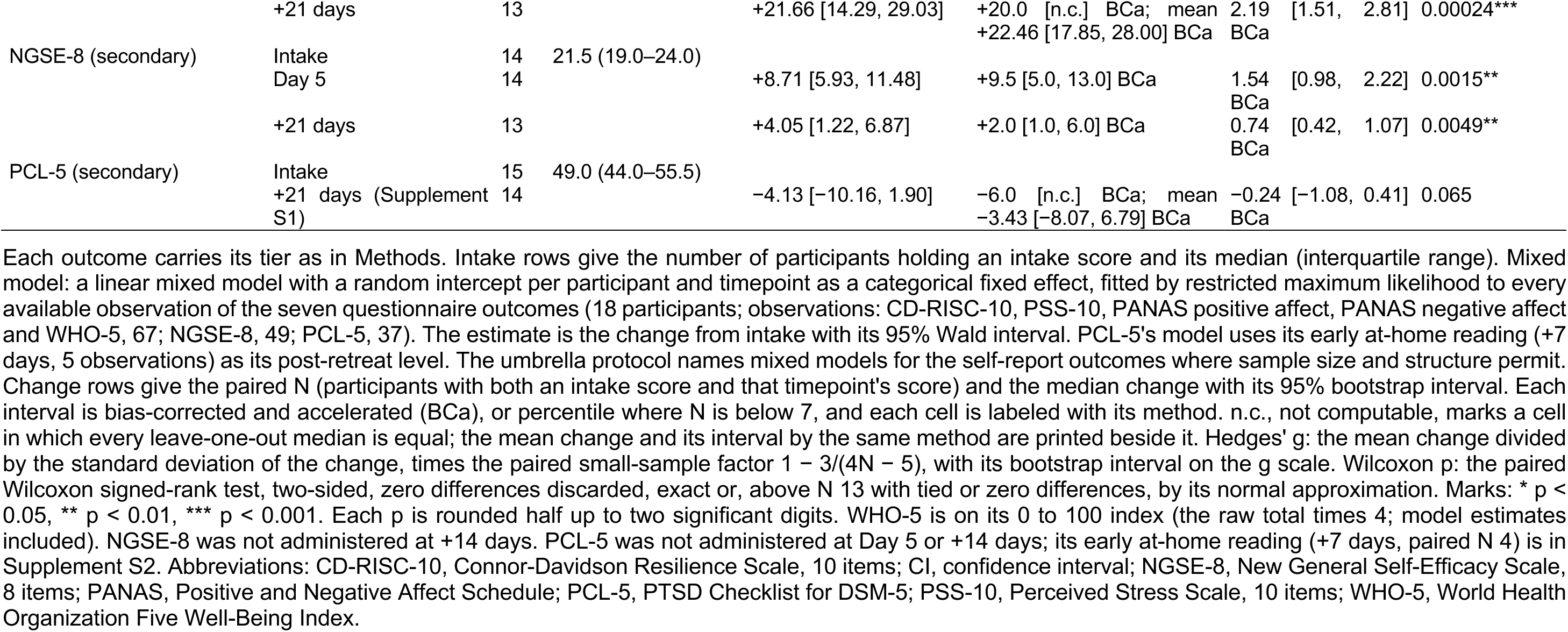
Self-report outcomes at intake, and change from intake at the end of the retreat (Day 5) and at +14 and +21 days.

### 2.3 Intervention

The intervention was a five-day, four-night residential nature-based resilience retreat in Finland, delivered by facilitators of the operator, Amoreh Oy. The program combined guided nature exposure, resonance-frequency paced breathing training, mindfulness training and meditation practice sessions along with moderate physical activity and group reflection sessions, with optional thermal contrast (sauna and cold-water immersion). Participants were asked to be phone-free during sessions, with brief in-app check-ins at session boundaries. The program by day and the app’s schedule are given in Supplement S6.

### 2.4 Measures

Questionnaires were administered in a dedicated companion app, offered in Hebrew and English, at intake, upon retreat completion (Day 5), and 14 and 21 days post-retreat. The licensed Hebrew CD-RISC-10 was used throughout. All 18 participants completed the Day-5 battery in the app (Supplement S3). On retreat days, the app preceded each self-report assessment except the sleep diary with a guided paced-breathing exercise that could not be skipped. A visual pacer set 26 breaths at 6 breaths per minute (5 s in, 5 s out; 4 min 20 s), with brief on-screen guidance instructions with no sound.

The primary outcome, the CD-RISC-10 questionnaire (Campbell-Sills and Stein, 2007; Connor and Davidson, 2003), has 10 items scored 0 to 4 (total 0 to 40) over the past month. The secondary outcomes were the Perceived Stress Scale test (PSS-10; Cohen and Williamson, 1988): 10 items, 0 to 4, items 4, 5, 7 and 8 reversed; total 0 to 40; past 7 days. The Positive and Negative Affect Schedule test (PANAS; Watson et al., 1988): 20 items, 1 to 5, momentary; positive and negative affect 10 to 50 each. The PTSD Checklist test for DSM-5 (PCL-5; Blevins et al., 2015; Bovin et al., 2016): 20 items, 0 to 4; total 0 to 80; past month. It was administered at intake, at home about 7 days after the retreat, and at +21 days. In this study, the PCL-5 was utilized as a symptom characterization instrument and not as participant screening. The Well-Being Index test (WHO-5; Topp et al., 2015): 5 items, 0 to 5, past two weeks; scaled 0 to 100. The New General Self-Efficacy Scale test (NGSE-8; Chen et al., 2001): 8 items, 1 to 5; total 8 to 40 points.

The exploratory outcome, the autobiographical future-thinking task (AFTT; optional), comprised five cued future-event narratives typed at intake and the last day of the retreat (Day 5). They were scored for episodic specificity, the proportion of details that are internal (specific to the imagined event), under the Autobiographical Interview classification (Levine et al., 2002) adapted for future events (Addis et al., 2008). Its rating, the second rater’s agreement and its result are detailed in Supplement S2.

At 14 and 21 days, participants who did not complete the scheduled in-app battery received the same closed-response items through a linked question sheet and returned their answers by message, for the reasons given in Supplement S3. Both routes were self-administered. Mood check-ins and sleep diaries are reported descriptively (Supplement S5).

Physiological data were collected alongside the questionnaires and are reported in a separate companion paper. The registered on-site physiological primary reported no change (N = 10; 95% BCa CI −0.20 to +0.03), informing the plan to measure whole-night heart rate variability at home after the next implementation.

### 2.5 Statistical analysis

To make use of every available measurement, change from intake was calculated for each battery instrument (the PANAS by subscale) with a linear mixed-effects model. The model had timepoint as a categorical fixed factor (with intake as reference) and a random intercept per participant. It was fitted by restricted maximum likelihood to every available observation at four timepoints (the NGSE-8 without +14; the PCL-5 at intake, the early at-home reading and +21), with Wald 95% CIs and p values shown. Two sensitivity analyses of the same model are reported in Supplement S4. In the first, each timepoint contrast was tested against a t distribution instead of the normal reference, with the between-within degrees of freedom: the number of observations minus the number of participants minus the number of timepoint terms (the small-sample reference). In the second, because the model assumes that observations are missing at random given the participant, the model was refitted on the participants who had an intake reading of the instrument. The registered family analysis is reported, with within-family false-discovery-rate control, in Supplement S2.

Paired contrasts against intake used every complete pair. They reported the median change with a bias-corrected and accelerated (BCa) bootstrap 95% CI (DiCiccio and Efron, 1996; Efron, 1987) (10,000 resamples, seed 20260809) with the percentile bootstrap in place of BCa in cells with N below 7, as registered. They also reported Cohen’s *d*_z_ (Lakens, 2013) with Hedges’ g and paired small-sample factor J = 1 − 3/(4N − 5) (Hedges, 1981). Its interval was taken over the resamples in which g could be computed and not reported when more than 1% failed (in the forest plot, the *d*_z_ interval is the g interval divided by the same factor). They used two-sided Wilcoxon signed-rank test, zero differences discarded. Where a median interval is not computable, the mean change and its BCa interval are reported beside it. Wilcoxon p below 0.05, 0.01 and 0.001 is marked *, ** and ***. To test the robustness of the results, we calculated the largest Cook’s distance of each contrast and reported it against the screen of 4/N. No participant was removed.

The OSF plan’s pre-registered secondary analysis (Supplement S2) groups CD-RISC-10 and PCL-5 at +21 and PSS-10 and PANAS at Day 5 under Benjamini-Hochberg false-discovery-rate control at q < 0.10 (Benjamini and Hochberg, 1995), together with AFTT specificity as a second family and leave-one-subject-out sensitivity.

### 2.6 Ethics and declarations

This study was approved by the Ben-Gurion University of the Negev Human Subjects Research Committee under the umbrella protocol (#1295-3). All participants in this study signed written informed consent before any data were collected. No adverse event was reported.

## 3. Results

### 3.1 Participants and follow-up

All 18 participants provided an end-of-retreat assessment. At 14 days post-retreat, all 18 provided follow-up data; at 21 days, 17 did so (Fig. 1). Baseline availability and paired sample sizes varied by instrument (Table 1; Supplement S3).

**Fig. 1.**
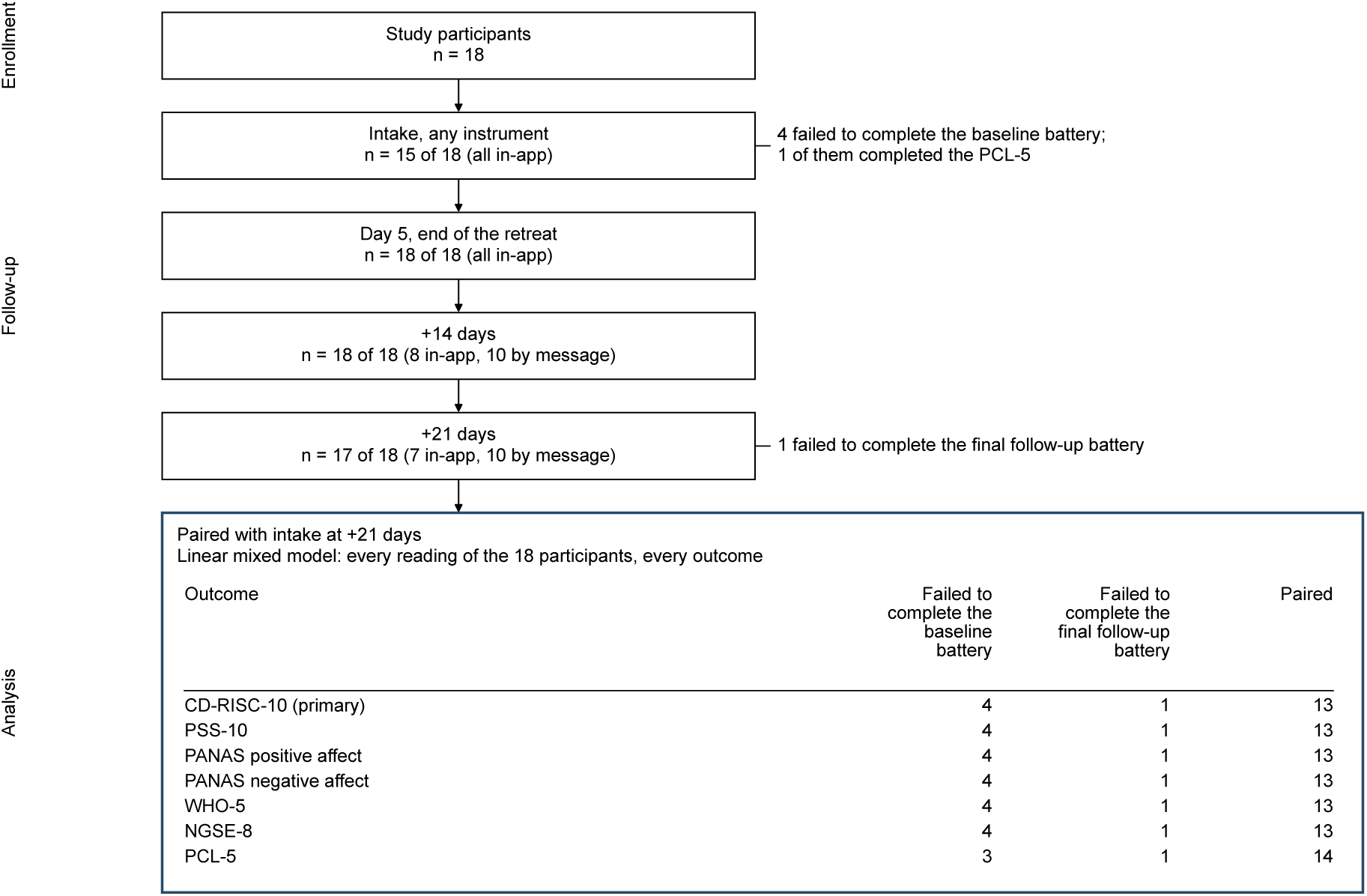
Participant flow for the self-report outcomes, from the N = 18 study participants. Each timepoint shows the number of participants with a reading there: intake and Day 5 in the companion app; +14 and +21 days in the app, or from a linked question sheet with answers returned by message where the app route was not completed. The analysis table gives, per outcome, the losses by cause and the participants paired with intake at +21 days. 4 failed to complete the baseline battery; 1 of them completed the PCL-5, so 3 lack a PCL-5 intake reading. 1 failed to complete the final follow-up battery. The mixed model uses every reading. The analysis step is outlined; no test is shown. Abbreviations: CD-RISC-10, Connor-Davidson Resilience Scale, 10 items; NGSE-8, New General Self-Efficacy Scale, 8 items; PANAS, Positive and Negative Affect Schedule; PCL-5, PTSD Checklist for DSM-5; PSS-10, Perceived Stress Scale, 10 items; WHO-5, World Health Organization Five Well-Being Index.

### 3.2 Primary outcome

From intake to +21 days post-retreat, the CD-RISC-10 change in the linear mixed model over the four timepoints was +5.08 points (95% Wald CI 2.51 to 7.65; p = 0.00011; 67 observations from 18 participants), with p = 0.00034 on the small-sample reference and +4.59 points (p = 0.0014) among the 14 participants with a complete baseline reading (Table S4b; Table S4d). In the paired contrast, the median CD-RISC-10 change was +4.0 points (N = 13; 95% BCa CI 0.0 to 7.0; Cohen’s *d*_z_ = 0.83; Hedges’ g = 0.78, 95% BCa CI 0.31 to 1.26; Wilcoxon p = 0.0098, q = 0.012 in the registered family; Table 1; Fig. 2A). One participant exceeded the pre-declared Cook’s distance screen of 4/N and was retained. Every leave-one-subject-out fold kept p below 0.05 (with the largest being 0.020; Supplement S2).

**Fig. 2.**
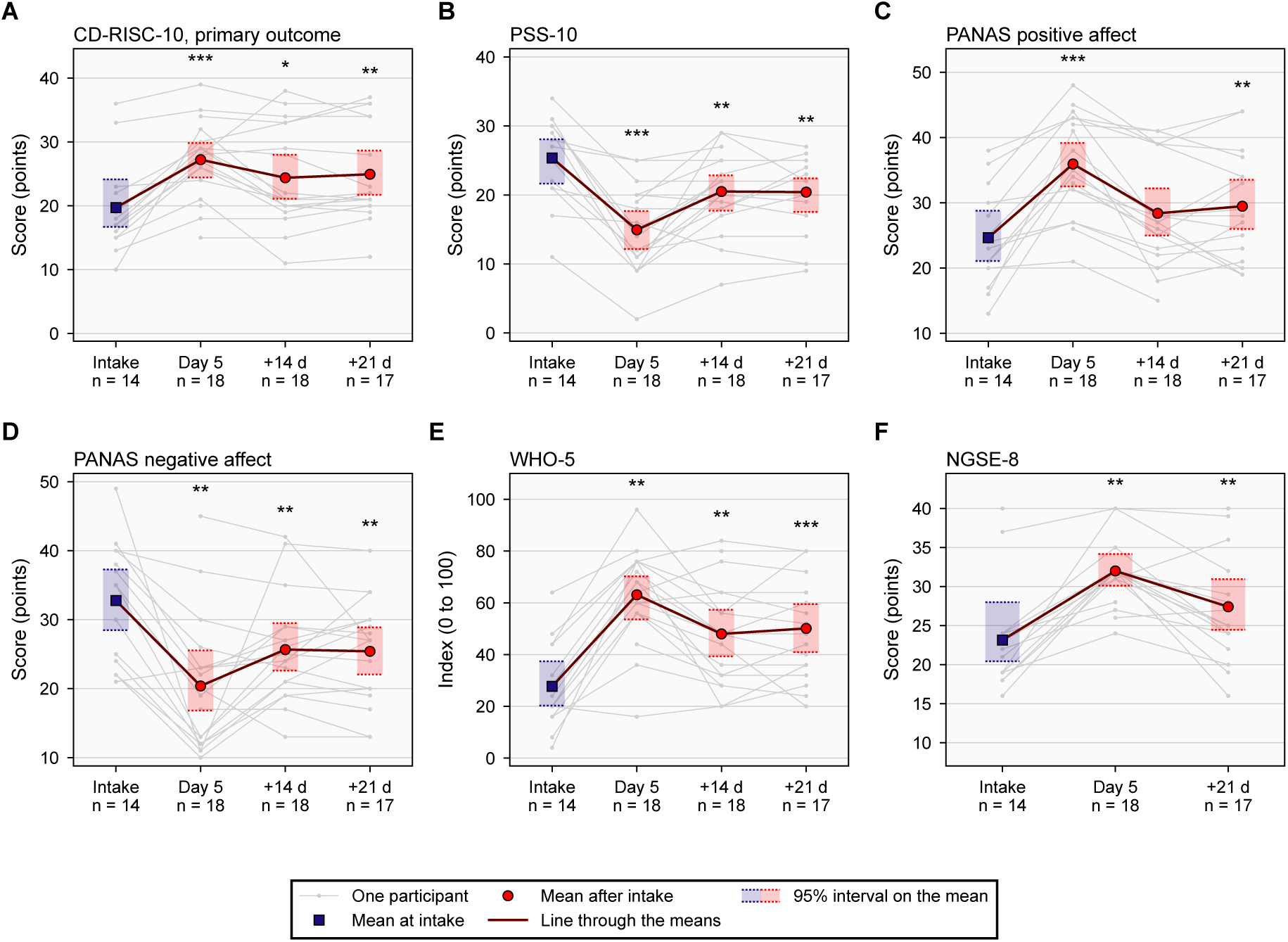
Self-report outcomes from intake to 21 days after the retreat. Each panel is one instrument on its full score range: CD-RISC-10, the primary outcome (A), PSS-10 (B), PANAS positive affect (C) and negative affect (D), WHO-5 on its 0 to 100 index (E) and NGSE-8 (F). NGSE-8 was not administered at +14 days. Gray lines join each participant’s readings. The square is the mean at intake and the circles the means after it, each over every reading at its timepoint, with its 95% interval on the mean shaded between dotted bounds (BCa bootstrap). A line joins the means. Each timepoint label shows the number of participants with a reading there, of the N = 18 participants analyzed. Asterisks mark the paired Wilcoxon signed-rank test of each timepoint against intake, two-sided: * p < 0.05, ** p < 0.01, *** p < 0.001. The paired N, median change, effect sizes and p for every timepoint are in Table 1; at +21 days the mixed model’s estimate and the paired contrast are both given there and in Results 3.2. Abbreviations: BCa, bias-corrected and accelerated; CD-RISC-10, Connor-Davidson Resilience Scale, 10 items; NGSE-8, New General Self-Efficacy Scale, 8 items; PANAS, Positive and Negative Affect Schedule; PSS-10, Perceived Stress Scale, 10 items; WHO-5, World Health Organization Five Well-Being Index.

### 3.3 Secondary outcomes

All six outcome measures were assessed at both Day 5 and 21 days. Paired changes from baseline persisted to day 21 post-retreat (each unadjusted Wilcoxon p ≤ 0.0098), although changes in score units were smaller than at Day 5 (Table 1; Fig. 2). Perceived stress and negative affect were lower, and positive affect, well-being and self-efficacy scores higher than baseline. In the mixed model, the secondary changes at +21 days of the PSS-10, both PANAS scales, the WHO-5 and the NGSE-8, all but the PCL-5’s, kept p below 0.05 on the small-sample reference and among the participants with a baseline reading (Supplement S4; Fig. 3; Fig. 4). Notably, four members of the registered family, the CD-RISC-10, the PSS-10 and both PANAS scales, met its criterion under every single omission (Supplement S2).

**Fig. 3.**
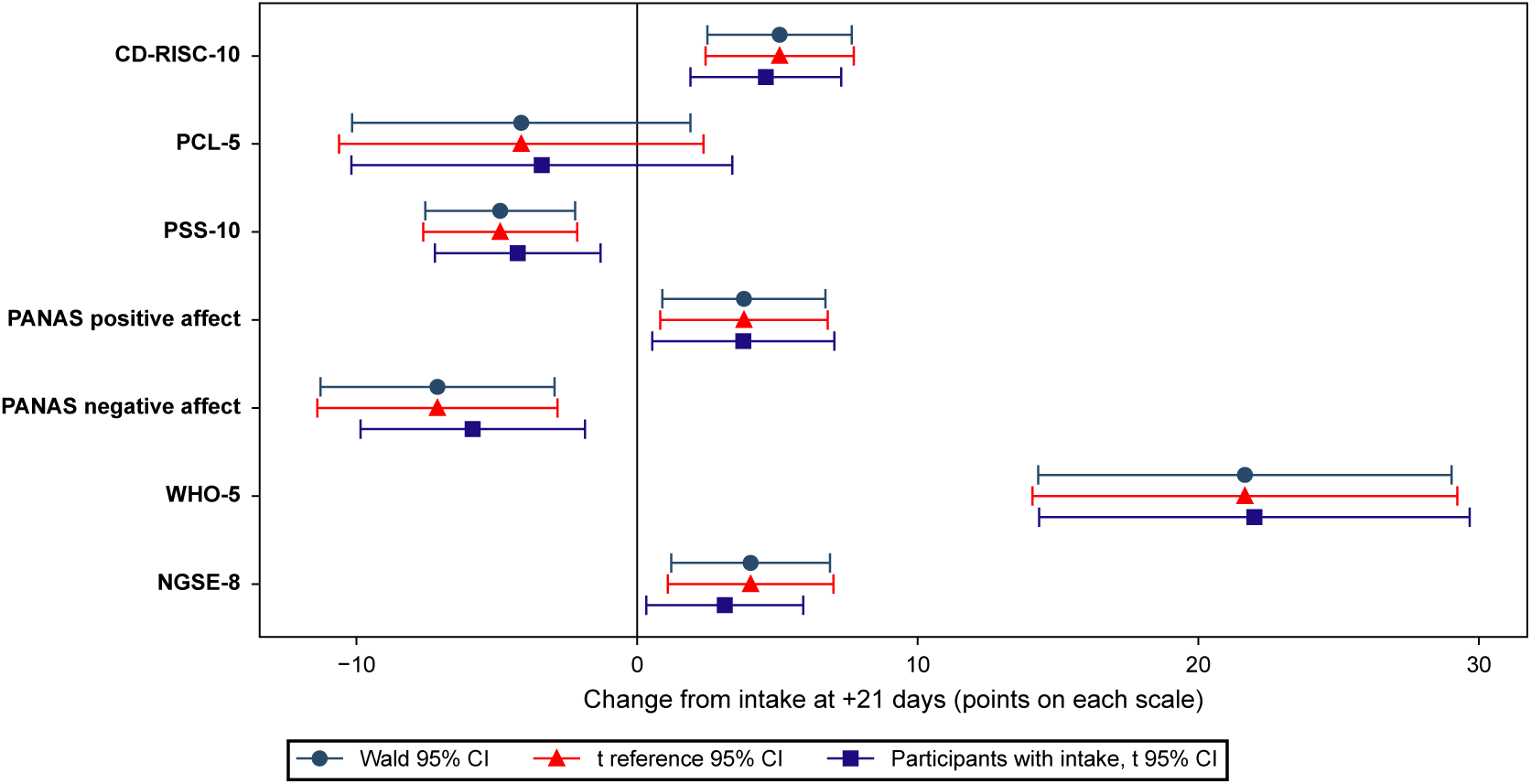
The +21-day change from intake under the mixed model and its two sensitivity analyses. Circles are the model’s estimate on every reading of the N = 18 participants with its Wald 95% interval. Triangles are the same estimate with its 95% interval on a t reference distribution. Squares are the model fitted on the participants with an intake reading (14, or 15 for the PCL-5), with its t interval. Each outcome is on its own scale, WHO-5 on its 0 to 100 index. The solid line marks zero. Every value is in Supplement S4. Abbreviations: CD-RISC-10, Connor-Davidson Resilience Scale, 10 items; NGSE-8, New General Self-Efficacy Scale, 8 items; PANAS, Positive and Negative Affect Schedule; PCL-5, PTSD Checklist for DSM-5; PSS-10, Perceived Stress Scale, 10 items; WHO-5, World Health Organization Five Well-Being Index.

**Fig. 4.**
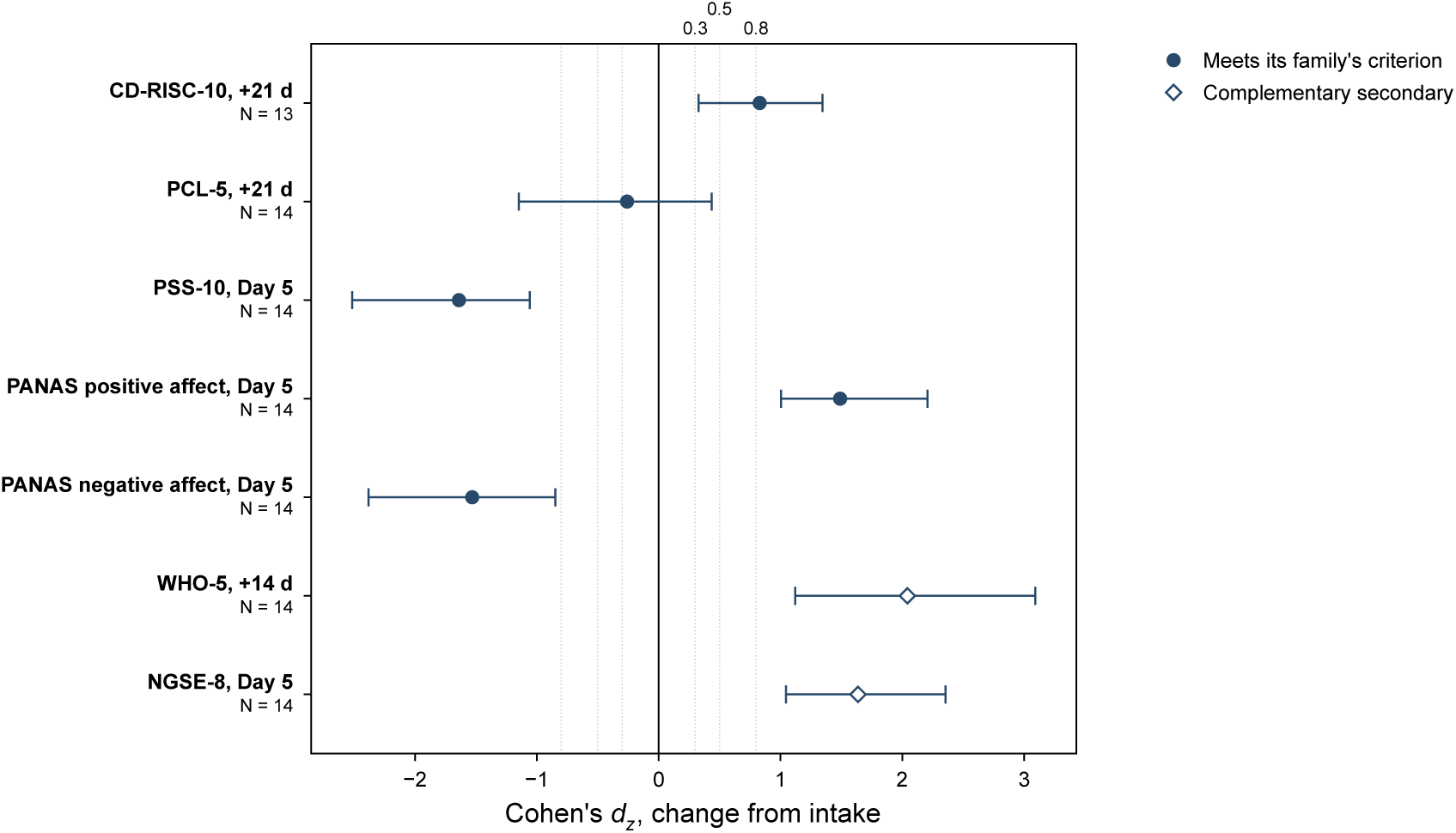
Change from intake on each member of the registered family at the timepoint of its registered contrast, and on the two complementary secondaries, WHO-5 at +14 days and NGSE-8 at Day 5, as each row names it. Each marker is Cohen’s *d*_z_ and each capped whisker its 95% interval, the bootstrap interval on Hedges’ g (Methods 2.5) divided by the row’s small-sample factor (BCa bootstrap). The PCL-5 row is the +21-day contrast (Supplement S1). Each row has N = 13 or N = 14 pairs. A filled circle marks a member that meets its family’s registered criterion (q < 0.10, Benjamini-Hochberg within the family; Supplement S2). Diamonds mark the two complementary secondaries, which belong to no family and have no verdict. Dotted guides mark the registration’s *d*_z_ thresholds of 0.3, 0.5 and 0.8 either side of the solid zero line. Abbreviations: BCa, bias-corrected and accelerated; CD-RISC-10, Connor-Davidson Resilience Scale, 10 items; NGSE-8, New General Self-Efficacy Scale, 8 items; PANAS, Positive and Negative Affect Schedule; PCL-5, PTSD Checklist for DSM-5; PSS-10, Perceived Stress Scale, 10 items; WHO-5, World Health Organization Five Well-Being Index.

The change in posttraumatic stress symptoms on the +21-day contrast (Supplement S1) was small and uncertain (PCL-5; N = 14; median change −6.0 points; Hedges’ g = −0.24, 95% BCa CI −1.08 to 0.41; Cohen’s *d*_z_ = −0.26; Wilcoxon p = 0.065). Applying the registered family’s correction gives q = 0.065, below the 0.10 family threshold, but the effect-size interval included zero, and single participant omissions moved p from 0.0059 to 0.11, with 1 of 14 crossing 0.05 (Supplement S2; Fig. 5).

**Fig. 5.**
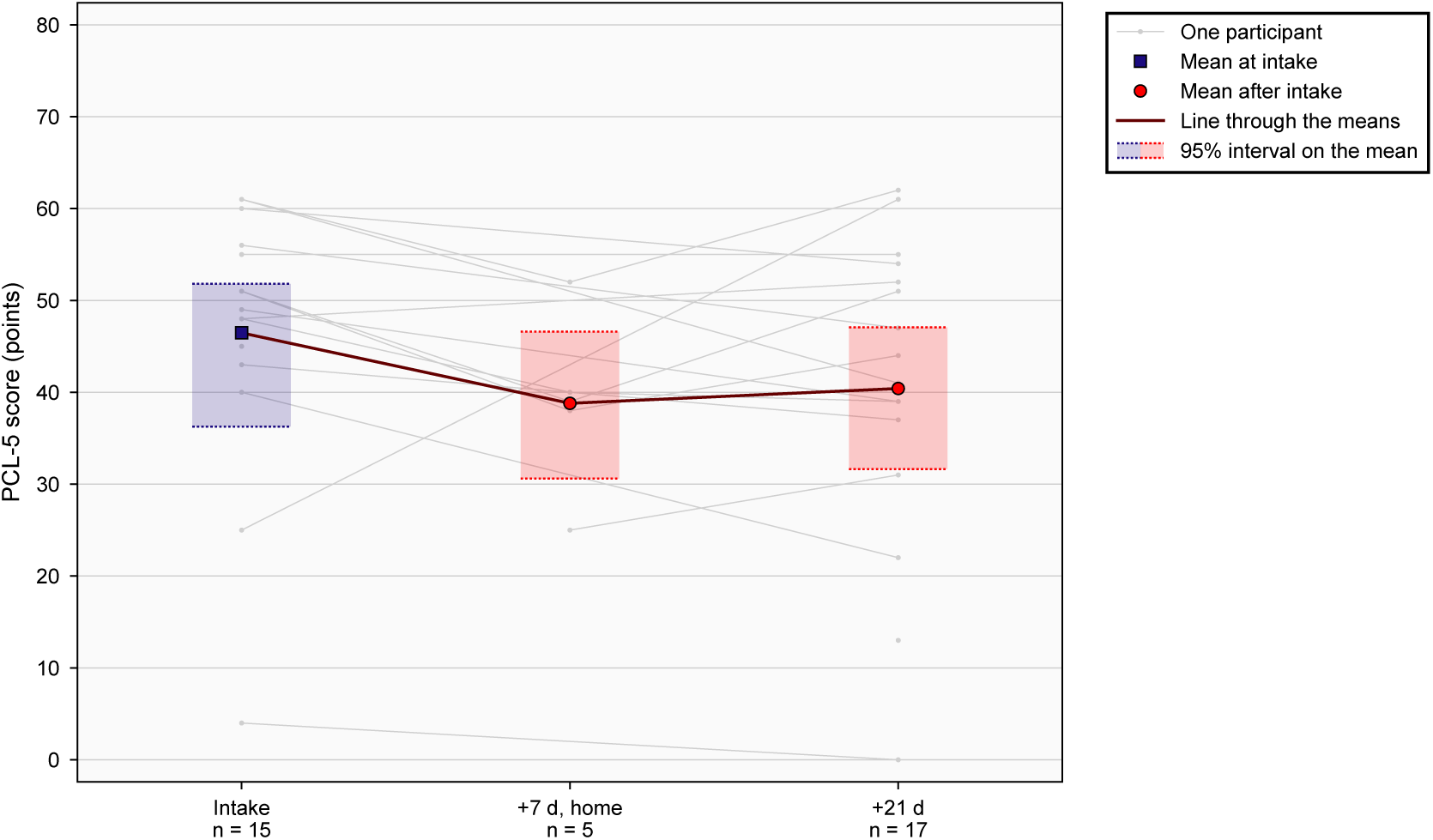
PCL-5 from intake to 21 days after the retreat. Gray lines join each participant’s readings. The square is the mean at intake and the circles the means after it, each over every reading at its timepoint, with its 95% interval on the mean shaded between dotted bounds (BCa bootstrap, or the percentile bootstrap for the +7-day timepoint, which has 5 readings). A line joins the means. The early at-home reading, a median of 7 days after the retreat, is an interim reading (Supplement S2). Each timepoint label shows the number of participants with a reading there, of the N = 18 participants analyzed. The change from intake to +21 days is tested in Results 3.3 (paired Wilcoxon signed-rank test, N = 14, W = 15.5, p = 0.065). Abbreviations: BCa, bias-corrected and accelerated; PCL-5, PTSD Checklist for DSM-5.

The median change in AFTT specificity, the exploratory measurement, from intake to the last day of the retreat was +0.092 (N = 6; 95% percentile CI −0.008 to 0.195; p = 0.16; Supplement S2; Fig. S1B).

## 4. Discussion

### 4.1 Principal findings

In this study we have found the resilience score was higher at 21 days post-retreat than at baseline. The protocol’s mixed model estimated a change of +5.08 points on the CD-RISC-10 (95% Wald CI 2.51 to 7.65; p = 0.00011; 67 observations from 18 participants). Perceived stress, positive and negative affect, well-being and general self-efficacy had also improved at +21 days (Table 1). For each of these scores the change in score points was largest upon retreat completion, partly subsided by +14 days post-retreat where measured, but with sustained improvement to +21 days post-retreat compared to baseline.

The evidence for improvement in posttraumatic stress symptoms was less decisive. The effect was small with an interval that included zero, although the result passed the 10% threshold. Nevertheless, the principal signal in this cohort applies to resilience and broader psychological well-being, rather than claiming amelioration of PTSD symptoms in particular.

### 4.2 Comparison with previous studies

Direct benchmarks for CD-RISC-10 change are limited. A randomized resilience trial in adolescents and young adults with cancer reported a 3.0-point benefit over usual care at six months (Rosenberg et al., 2018). A four-week heart rate variability biofeedback program also improved resilience relative to a wait-list, with follow-up maintenance (Cortez-Neavel et al., 2026). These controlled estimates cannot be directly compared with the within-group changes reported (Morris and DeShon, 2002) and therefore do not claim a clinical pertinence threshold for this cohort. A closer design analog, an uncontrolled week-long wellness retreat, reported improvements maintained at six weeks among participants followed up (Cohen et al., 2017).

Follow-up improvements have also been reported after related programs. Studies with recreational intervention comparison groups illustrate that improvement during a residential stay can occur in both groups, highlighting the importance of a comparator when attributing change to specific program components (Epel et al., 2016), as in one trial reported twice (Hwang et al., 2018; Kwak et al., 2019). Meta-analyses report maintained gains following meditation retreats (Khoury et al., 2017) and partial persistence after positive-psychology interventions (Carr et al., 2024).

### 4.3 Strengths and limitations

The strengths of this study include longitudinal follow-up through 21 days post-retreat, high follow-up completion, and rigorous reporting of individual trajectories and effect sizes with uncertainty intervals with its secondary analysis publicly pre-registered. Nevertheless, this study has several limitations: the absence of a comparison group and the small sample size are chief among these. Age and sex were not linked to the research dataset, to preserve participants’ privacy in a small group, which limits generalizability. The questionnaire administration route varied: where some participants could not complete the follow-up batteries through the app, a message-based follow-up route was served. Four end-of-retreat assessments were delayed due to return voyage connectivity issues and Shabbat considerations. The study measured perceived resilience, not subsequent functioning under a new stressor. The Hebrew CD-RISC-10 has previously been used in an Israeli sample of 161 physicians, in which it demonstrated good internal consistency (Cronbach’s α = 0.869; Joachim et al., 2024). The findings in this study are pre-registered as hypothesis-generating.

### 4.4 Implications

The findings compel controlled evaluation of nature-based residential programs. The partial attenuation after return to routine supports testing post-retreat impact and follow-up. A future study could compare participants with a prospectively defined wait-list, with allocation and assessment procedures designed to limit selection bias. Standardized assessment conditions, documented activity and sleep, and resting or home-night physiological measurements would help distinguish psychological change from short-term physical load.

### 4.5 Conclusions

In this cohort, self-reported resilience remained higher three weeks after a five-day nature-based retreat, alongside lower perceived stress and negative affect and higher positive affect, well-being and self-efficacy. These consistent within-group changes support a prospective controlled evaluation.

## Declaration of AI-assisted technologies

During the preparation of this work the authors used AI tools to improve readability and language of this manuscript.

## Author contributions: CRediT

**Raz Balin:** Conceptualization, Methodology, Software development, Validation, Statistical analysis, Investigation, Data curation, Visualization, Project administration, Writing – original draft, Writing – review and editing.

**Matan Lev:** Data analysis.

**Adi Schneider Moreh:** Design and delivery of the intervention.

**Itamar Grotto:** Conceptualization, Methodology, Supervision, Project administration, Writing – review and editing.

## Data Availability

The coded dataset contains sensitive health information from a small cohort and is therefore not publicly deposited due to risk of participant identification and privacy considerations. Access may be requested from the corresponding author under the data controller’s access arrangements at Ben-Gurion University of the Negev. Analysis code is available from the corresponding author upon request. The registration and posted amendments are publicly available at https://osf.io/7wkm5.

## Acknowledgments

We thank the participants.

