## Supplementary material for "Resilience following a nature-based retreat: a prospective observational study"

S1, deviations from the registered plan; S2, the registered secondary analysis, with the exploratory AFTT outcome; S3, the sample-size cascade by cause; S4, the mixed-effects model and the sensitivity analyses; S5, descriptive figures; S6, the retreat program.

**Conventions.** N is the number of participants with both readings of a contrast; a change is the later reading minus the earlier. Intervals are 95% bootstrap intervals (10,000 resamples, seed 20260809, except where stated for rater-agreement intervals), bias-corrected and accelerated (BCa) except in cells with N below 7, which take the percentile bootstrap as registered (section 6.5 of the registration). Every interval cell is labeled with its method. n.c. (not computable) is followed by its reason in the cell or explained in the note to its table. p is the two-sided paired Wilcoxon signed-rank p with zero differences discarded, exact where the differences carry no ties or zeros, exact by full permutation up to N 13 where they do, and by the normal approximation above N 13. q is the Benjamini-Hochberg false-discovery-rate value within a registered family, with rejection at q < 0.10. Hedges' g carries the paired small-sample factor. For a leave-one-subject-out (LOSO) row, each participant is omitted in turn and the test repeated; it gives the range of p over the omissions and the omissions whose p lies on the other side of 0.05 from the full sample's p. Cook's d is the largest Cook's distance of the change scores against the pre-declared screen of 4/N, with the participants above it in parentheses, all retained. Values are rounded half up.

#### S1. Deviations from the registered plan

S1 lists the deviations cited in this paper and supplement, using the numbering of the analysis record. Amendments 1 to 6 are posted at osf.io/7wkm5.

**Table S1.** Deviations cited in this paper.

| Deviation | Date | What it changed | Registration record |
|---|---|---|---|
| 4 | 2026-06-30 | PCL-5 administered at most three times, at home only: intake, an early post about 7 to 10 days after the retreat, and +21 | Amendment 1 |
| 5 | 2026-07-02 | PCL-5 recall window harmonized to the validated past month at all three at-home administrations | Amendment 1 |
| 6 | 2026-07-02 | PSS-10 recall stem set to the past 7 days for repeated administration, pre-onset | Amendment 1 |
| 7 | 2026-07-04; co-signed 2026-08-18 | long-window instruments read at +21 (CD-RISC-10) and +14 (WHO-5), the Day-5 readings kept as interim | Amendment 1 |
| 9 | 2026-07-04 | NGSE-8 reduced to three administrations; +14 dropped | Amendment 1 |
| 19 | 2026-07-24 | the CD-RISC-10 rendered in its licensed Hebrew in every battery | Amendment 3 |
| 29 | 2026-07-28 | PCL-5 administered four times (the intake battery's PCL-5 added to the three at-home administrations); the at-home administration pre-declared as the intake of record | Amendment 6 |
| 31 | 2026-07-28 | The intake battery was self-administered in the app on the participant's own phone, at or after the in-person intake session, on both handset platforms; the setting of each administration was not recorded. The battery's consent task was presented in that setting. The Principal Investigator's determination: written consent was taken in person at the intake session, so the consent task in the app followed it and was included in the approved procedure. | Amendment 6 |
| 32 | 2026-08-08 | the retreat-day breathing practice compelled without a skip control before every retreat-day session except the sleep diary; practice-abandonment named as a missingness mechanism | Amendment 4 |

| Deviation | Date | What it changed | Registration record |
|---|---|---|---|
| 33 | 2026-08-08 | every affect rating on a retreat day was made after the app's compelled practice; the confound declared with two sensitivities | Amendment 4 |
| 39 | 2026-08-08 | the PANAS intake of record is the at-home anchor where both administrations exist, parallel to Deviation 29 | Amendment 5 |
| 53 | 2026-08-19 | a third baseline provenance category (the late battery of retreat Day 1); the intake battery of four study participants is missing, one of whom completed the at-home PCL-5 intake | Amendment 6 |
| 54 | 2026-08-21 | AFTT rubric of record re-anchored to the Autobiographical Interview classification as adapted for future events | Amendment 6 |
| 55 | 2026-08-21 | narrative length registered as a residual blinding channel; second-rater agreement fixed as a per-kind ICC(A,1), prevalence-weighted, with the registered 0.70 threshold on the aggregate | Amendment 6 |
| 56 | 2026-08-26 | the first AFTT rating pass withdrawn because the rubric was incomplete when it was applied; the pass of record is the one rated under the completed rubric | Amendment 6 |
| 57 | 2026-08-27 | 10 study participants completed the +14 battery by an alternative route; mode recorded per row | Amendment 6 |
| 58 | 2026-08-28 | the Hebrew CD-RISC-10 instruction in the app carried a literal English placeholder at every administration, as shipped, and the question sheets of +14 and +21 did not; both routes named the past month; after the Hebrew sentence the placeholder read, in English and in angle brackets, 'official past-month HE instruction from the licensed PDF' | Amendment 6 |
| 63 | 2026-09-04 | the +21 alternative route contains the closed-response items only; modes and causes enumerated per participant (7 in-app, 10 alternative route, 1 lost to follow-up); the last reading on 2026-09-04; the +21 AFTT excluded for every participant | Amendment 6 |
| 64 | 2026-09-05 | data freeze declared 2026-09-05 instead of 2026-09-17; frozen analysis set fixed; paired PCL-5 N 14, above Amendment 5's floor of 4; F12's adherence counted under its registered rule | Amendment 6 |
| 66 | 2026-09-09; corrected 2026-09-28 | the PCL-5 contrast assessed at +21 days, the early post an interim reading outside every family, a reading of a posted record that is not uniform on the point, ruled on 2026-09-28 after the results were seen | Amendment 6 |
| 71 | 2026-09-26 | the confound declaration and sensitivity (ii) reported as registered | Amendment 6 |
| 72 | 2026-09-26 | the PANAS baseline provenance line derivable and reported | Amendment 6 |
| 73 | 2026-09-26 | the +14 and +21 alternative-route mode corrected to the route used (a linked question sheet, answers returned by message); the first author's transcription recorded against the registered blinding | this supplement |
| 74 | 2026-09-26 | Amendment 5's same-day admissibility clause not applied in the analysis of record (PANAS 4 of 14, AFTT 2 of 6 pairs late); the same-day restriction reported under Deviation 77 | Amendment 6 |
| 75 | 2026-09-26 | the registered covariates age and sex are held on the intake record apart from the research data and were not linked to the coded study dataset | this supplement |
| 77 | 2026-10-04 | three sensitivity analyses added on the principal investigator's review, their terms fixed before any was run: the Day-5 contrasts restricted to same-day pairs, the mixed model's timepoint contrasts read on a t reference with the between-within degrees of freedom, and that model fitted on the participants with an intake reading | this supplement |

| Deviation | Date | What it changed | Registration record |
|---|---|---|---|
| 78 | 2026-10-05 | One completed PANAS intake reading was restored from the frozen source sessions (N = 14). No other reading that enters an analysis changed. | this supplement |

### S2. The registered secondary analysis (osf.io/7wkm5)

The physiological-domain registration, frozen before any test was run, groups the CD-RISC-10, PCL-5, PSS-10 and the PANAS into family F9, analyzed at its registered contrasts, the PCL-5 at the +21-day contrast (Supplement S1), with the Wilcoxon signed-rank test under Benjamini-Hochberg control within the family. The AFTT forms family F10. WHO-5 and NGSE-8 are registered complementary secondaries outside every family. The registration admits a Day-5 PANAS or AFTT record only when taken on the same study-local day (Amendment 5, Clause 2). The analysis of record did not apply that rule, so the PANAS rows include 4 of 14 pairs read 1 to 4 days late and the AFTT row 2 of 6 read 1 to 2 days late (Deviation 74). Restricted to same-day pairs as a sensitivity (Deviation 77), the Day-5 contrasts are in Table S4c.

**Classification rule.** A family member is marked "rejects" when its q, computed within its own family, is below 0.10; WHO-5 and NGSE-8 have no q and no verdict. The $d_z$ band is the registration's frozen interpretation band (section 6.4: large from 0.8, moderate from 0.5, modest from 0.3, negligible below 0.3). For the PCL-5 the table gives the +21-day contrast (Supplement S1) and, as an interim row outside every family, the early at-home reading.

**Table S2.** The registered behavioral families at their registered contrasts, with the two complementary secondaries and the PCL-5 early at-home reading.

| Outcome | Family | Registered contrast | N | Median change [95% CI] | Mean change | $d_z$ (band) | Hedges' g [95% CI] | W | p | q | Verdict at q < 0.10 | LOSO p range (omissions) | Omissions crossing p = 0.05 | Cook's d max (above 4/N) |
|---|---|---|---|---|---|---|---|---|---|---|---|---|---|---|
| CD-RISC-10 | F9 | intake to +21 days | 13 | +4.0 [0.0, 7.0] BCa | +4.92 | 0.83 (large) | 0.78 [0.31, 1.26] BCa | 7 | 0.0098 | 0.012 | rejects | 0.0049 to 0.020 (13) | 0 of 13 | 0.44 (1) |
| PCL-5 | F9 | intake to +21 days (Supplement S1) | 14 | −6.0 [n.c.] BCa | −3.43 [−8.07, 6.79] BCa | −0.26 (negligible) | −0.24 [−1.08, 0.41] BCa | 15.5 | 0.065 | 0.065 | rejects | 0.0059 to 0.11 (14) | 1 of 14 | 0.74 (1) |
| PCL-5 | none | intake to +7 days, early at-home post (interim reading) | 4 | −10.5 [−13.0, −8.0] percentile | −10.50 | −4.41 (large) | −3.21 [n.c.: degenerate bootstrap] percentile | 0 | 0.13 | not applicable | interim reading; outside every family | 0.25 to 0.25 (4) | 0 of 4 | 0.49 (0) |
| PSS-10 | F9 | intake to Day 5 | 14 | −12.0 [n.c.] BCa | −11.21 [−14.57, −7.64] BCa | −1.64 (large) | −1.54 [−2.37, −1.00] BCa | 0 | 0.00096 | 0.0020 | rejects | 0.00024 to 0.00024 (14) | 0 of 14 | 0.18 (0) |
| PANAS positive affect | F9 | intake to Day 5 | 14 | +13.0 [n.c.] BCa | +11.21 [7.57, 15.21] BCa | 1.49 (large) | 1.40 [0.95, 2.08] BCa | 0 | 0.00098 | 0.0020 | rejects | 0.00024 to 0.00024 (14) | 0 of 14 | 0.28 (0) |
| PANAS negative affect | F9 | intake to Day 5 | 14 | −12.5 [−17.0, −10.0] BCa | −14.07 | −1.53 (large) | −1.44 [−2.24, −0.80] BCa | 1 | 0.0012 | 0.0020 | rejects | 0.00024 to 0.00049 (14) | 0 of 14 | 0.56 (1) |

| Outcome | Family | Registered contrast | N | Median change [95% CI] | Mean change | $d_z$ (band) | Hedges' g [95% CI] | W | p | q | Verdict at q < 0.10 | LOSO p range (omissions) | Omissions crossing p = 0.05 | Cook's d max (above 4/N) |
|---|---|---|---|---|---|---|---|---|---|---|---|---|---|---|
| AFTT specificity | F10 | intake to Day 5 | 6 | +0.092 [−0.008, 0.195] percentile | +0.0933 | 0.88 (large) | 0.74 [0.09, 3.04] percentile | 3 | 0.16 | 0.16 | does not reject | 0.063 to 0.31 (6) | 0 of 6 | 0.73 (1) |
| WHO-5 (0 to 100 index) | none | intake to +14 days | 14 | +20.0 [n.c.] BCa | +18.57 [13.71, 23.14] BCa | 2.04 (large) | 1.92 [1.05, 2.91] BCa | 0 | 0.0014 | not applicable | registered complementary secondary; outside every family; not corrected | 0.00024 to 0.00049 (14) | 0 of 14 | 0.35 (2) |
| NGSE-8 | none | intake to Day 5 | 14 | +9.5 [5.0, 13.0] BCa | +9.43 | 1.64 (large) | 1.54 [0.98, 2.22] BCa | 0 | 0.0015 | not applicable | registered complementary secondary; outside every family; not corrected | 0.00024 to 0.00049 (14) | 0 of 14 | 0.33 (1) |

The registration's exploratory sign-aware Stouffer composite combines the members of F9 (CD-RISC-10, PCL-5, PSS-10, PANAS positive affect and PANAS negative affect), PCL-5 at the +21-day contrast (Supplement S1): z 6.38, p $1.8 \times 10^{-10}$. Every self-report completed on a retreat day except the sleep diary followed the app's compelled breathing practice, and the intake readings did not; for the PANAS that difference works in the direction of improvement (Deviations 32 and 33). Table 1 gives the PANAS at +21 days and its sensitivity (ii). The median intervals marked n.c. (PCL-5, PSS-10, PANAS positive affect and WHO-5) are not computable because every leave-one-out median is equal; the mean change and its interval are printed beside them.

The exploratory outcome, AFTT specificity, forms family F10. The first author rated the narratives blind to timepoint. A second rater double-rated 33 of the 130 narratives, with agreement (per-kind intraclass correlation ICC(A,1), prevalence-weighted over the 7 of 10 estimable detail kinds) of 0.87 (95% BCa CI 0.72 to 0.97, 10,000 resamples) against the registered threshold of 0.70 (Deviation 55). The median change from intake to Day 5 was +0.092 (N = 6; 95% percentile CI −0.008 to 0.195; p = 0.16; Fig. S1). The score is the proportion of internal details among all details, and Day-5 narratives contained fewer details than intake narratives, so the median change cannot be separated from narrative length. The registration's declared tertiary AFTT perceptual to thought ratio, Day 5 against intake, is exploratory, untested and uncorrected: N = 6, median change −0.129 (95% percentile CI −0.350 to 0.044), Cohen's $d_z$ = −0.77.

The same registration's physiological families are the subject of a companion measurement paper. Two behavioral families were not delivered. F12, in-app adherence and daily check-ins: 2 participants met its registered adherence rule of 11 or more of 21 days (1 without substitute completions; Deviation 64), below the family floor of 4. As Amendment 5 Clause 6 registers for a family not delivered, it is reported as not delivered, never as a finding of no effect. Its adherence distribution, the days with a completed check-in in the 21-day window, substitute completions counted (Deviation 64): none, 5; 1 to 5, 8; 6 to 10, 3; 11 or more, meeting the registered rule, 2. F13, the follow-up free-text family: its +21 future-thinking administration was excluded for every participant (Deviation 63).

#### S3. The sample-size cascade, by cause at every gate

Counts in Tables S3a to S3c are of study participants unless a row is labeled otherwise. Each loss is listed with its cause where one is recorded; a loss of the same participants from several instruments is listed under each.

**Table S3a.** Participants at each gate of the behavioral assessments.

| Gate | N | Losses by cause, and the route of administration |
|---|---|---|
| Study identifiers issued (Amendment 5) | 20 identifiers | 1 never paired, a duplicate enrollment of a participant who re-enrolled; 1 held by the retreat operator, outside the analysis set |
| Study participants | 18 | all analyzed |
| Intake battery (CD-RISC-10, PSS-10, WHO-5, NGSE-8) | 14 | 4 failed to complete the baseline battery (Deviation 53): 3 with no intake reading of any instrument, 1 with the at-home PCL-5 only |
| Intake PANAS | 14 | the same 4 |
| Intake PCL-5 | 15 | 3 of the same 4; the remaining 1 completed the PCL-5 intake by its separate at-home route |
| Intake AFTT | 6 | the same 4; 8 further participants with no intake narrative set |
| Day 5 (end of the retreat) | 18 | none lost; 18 in the app; 14 on the last retreat day and 4 later, at most 4 days after it |
| +14 days | 18 | none lost; 8 in the app; 10 from a linked question sheet with answers returned by message (Deviations 57 and 73): 6 had stopped submitting in the app, 4 had completed another in-app session in its place; 14 have both the intake battery and +14, 18 have both Day 5 and +14 |
| +21 days | 17 | 1 lost to follow-up (Deviation 63); 7 in the app; 10 from a linked question sheet with answers returned by message (Deviations 63 and 73): handset lost 2, app deleted 2, app defect 3, window missed 3 |

**Table S3b.** Reach of the registered physiological primary outcome, whose endpoints are the first and the last retreat day (the outcome itself is reported in this paper's Methods).

| Gate | Participants | Not reaching the gate |
|---|---|---|
| A Day-1 value | 15 | 3 |
| A Day-5 value | 11 | 7 |
| Both endpoints (the analyzed N) | 10 | 8, by admission pattern: Day 1 reached, Day 5 not reached (no data in the session window), 5; 3 in patterns each shared by fewer than 3, merged: Day 1 not reached (zero usable pulse beats or no data in the session window), Day 5 not reached or reached (no data in the session window) |

The endpoint values not reached, by cause: no data in the session window 9; zero usable pulse beats 1.

**Table S3c.** The registered cascade lines on intake provenance and on the clocks that time a reading.

| Line | N | Cause or treatment |
|---|---|---|
| PANAS intake reading of record, by source (Deviation 39) | 14 | the at-home PANAS 3; the intake battery 11 (6 on 2026-07-29, 4 on 2026-08-02, 1 on retreat Day 1) |
| PANAS intake collisions resolved by rule (Deviation 39) | 8 | 7 by the Deviation 39 rule, the at-home anchor kept; 1 by the heavier-packet rule |
| Readings timed by the device's finalization clock (Amendment 5, Clause 4) | 119 readings from 7 participants | scored questionnaire readings at the four registered timepoints, each admitted and flagged; the device's clock recorded the capture time (Amendment 5 requires this clock to be counted per session) |

### S4. The mixed-effects model and the sensitivity analyses

**Table S4a.** The linear mixed-effects model of the Methods, fitted to every available observation: model summary.

| Outcome | Participants | Observations | Readings by timepoint |
|---|---|---|---|
| CD-RISC-10 | 18 | 67 | Intake 14; Day 5 18; +14 days 18; +21 days 17 |
| PCL-5 | 18 | 37 | Intake 15; early at-home post 5; +21 days 17 |
| PSS-10 | 18 | 67 | Intake 14; Day 5 18; +14 days 18; +21 days 17 |
| PANAS positive affect | 18 | 67 | Intake 14; Day 5 18; +14 days 18; +21 days 17 |
| PANAS negative affect | 18 | 67 | Intake 14; Day 5 18; +14 days 18; +21 days 17 |
| WHO-5 (0 to 100 index) | 18 | 67 | Intake 14; Day 5 18; +14 days 18; +21 days 17 |
| NGSE-8 | 18 | 49 | Intake 14; Day 5 18; +21 days 17 |

**Table S4b.** Every estimated change from intake of the models in Table S4a: the model's Wald interval and p and, as a sensitivity (Deviation 77), the interval and p on a t reference distribution whose degrees of freedom (df) are the observations minus the participants minus the timepoint terms. The estimate and its standard error are the model's. WHO-5's model is fitted on the raw 0 to 25 total; its estimate, standard error and interval are shown on the 0 to 100 index (times 4), as in Table 1, and its z and p are the model's.

| Outcome | Term | Estimate | SE | 95% CI | z | p | df | 95% CI (t) | p (t) |
|---|---|---|---|---|---|---|---|---|---|
| CD-RISC-10 | +21 days against intake | +5.08 | 1.31 | [2.51, 7.65] Wald | 3.87 | 0.00011 | 46 | [2.44, 7.72] t | 0.00034 |
| CD-RISC-10 | +14 days against intake | +4.59 | 1.29 | [2.06, 7.11] Wald | 3.56 | 0.00037 | 46 | [2.00, 7.18] t | 0.00087 |
| CD-RISC-10 | Day 5 against intake | +7.42 | 1.29 | [4.90, 9.94] Wald | 5.76 | $8.2 \times 10^{-9}$ | 46 | [4.83, 10.01] t | $6.5 \times 10^{-7}$ |
| PCL-5 | +21 days against intake | −4.13 | 3.08 | [−10.16, 1.90] Wald | −1.34 | 0.18 | 17 | [−10.62, 2.36] t | 0.20 |
| PCL-5 | Early at-home post against intake | −9.90 | 4.87 | [−19.44, −0.36] Wald | −2.03 | 0.042 | 17 | [−20.17, 0.37] t | 0.058 |
| PSS-10 | +21 days against intake | −4.88 | 1.36 | [−7.55, −2.21] Wald | −3.58 | 0.00034 | 46 | [−7.62, −2.14] t | 0.00082 |
| PSS-10 | +14 days against intake | −5.08 | 1.34 | [−7.70, −2.46] Wald | −3.80 | 0.00015 | 46 | [−7.77, −2.39] t | 0.00043 |
| PSS-10 | Day 5 against intake | −10.64 | 1.34 | [−13.26, −8.01] Wald | −7.95 | $1.8 \times 10^{-15}$ | 46 | [−13.33, −7.94] t | $3.5 \times 10^{-10}$ |
| PANAS positive affect | +21 days against intake | +3.81 | 1.48 | [0.90, 6.72] Wald | 2.57 | 0.010 | 46 | [0.82, 6.79] t | 0.014 |
| PANAS positive affect | +14 days against intake | +3.32 | 1.46 | [0.46, 6.17] Wald | 2.28 | 0.023 | 46 | [0.39, 6.24] t | 0.027 |
| PANAS positive affect | Day 5 against intake | +10.87 | 1.46 | [8.02, 13.72] Wald | 7.47 | $8.0 \times 10^{-14}$ | 46 | [7.94, 13.80] t | $1.8 \times 10^{-9}$ |
| PANAS negative affect | +21 days against intake | −7.12 | 2.13 | [−11.29, −2.95] Wald | −3.34 | 0.00082 | 46 | [−11.40, −2.83] t | 0.0016 |
| PANAS negative affect | +14 days against intake | −7.14 | 2.09 | [−11.23, −3.04] Wald | −3.42 | 0.00064 | 46 | [−11.34, −2.93] t | 0.0013 |
| PANAS negative affect | Day 5 against intake | −12.42 | 2.09 | [−16.51, −8.32] Wald | −5.94 | $2.8 \times 10^{-9}$ | 46 | [−16.62, −8.21] t | $3.6 \times 10^{-7}$ |
| WHO-5 (0 to 100 index) | +21 days against intake | +21.66 | 3.76 | [14.29, 29.03] Wald | 5.76 | $8.3 \times 10^{-9}$ | 46 | [14.09, 29.23] t | $6.6 \times 10^{-7}$ |
| WHO-5 (0 to 100 index) | +14 days against intake | +20.56 | 3.69 | [13.32, 27.79] Wald | 5.57 | $2.6 \times 10^{-8}$ | 46 | [13.13, 27.98] t | $1.3 \times 10^{-6}$ |
| WHO-5 (0 to 100 index) | Day 5 against intake | +35.67 | 3.69 | [28.43, 42.90] Wald | 9.66 | $4.3 \times 10^{-22}$ | 46 | [28.24, 43.10] t | $1.2 \times 10^{-12}$ |
| NGSE-8 | +21 days against intake | +4.05 | 1.44 | [1.22, 6.87] Wald | 2.80 | 0.0051 | 29 | [1.09, 7.00] t | 0.0089 |
| NGSE-8 | Day 5 against intake | +8.71 | 1.41 | [5.93, 11.48] Wald | 6.16 | $7.5 \times 10^{-10}$ | 29 | [5.81, 11.60] t | $1.0 \times 10^{-6}$ |

**Table S4c.** The registered Day-5 contrasts restricted to same-day pairs, a sensitivity (Deviation 77): each contrast on the pairs whose Day-5 reading was taken on the last retreat day, beside its row of record in Table S2. The family verdicts of record are unchanged, and the restricted rows have no q and no verdict.

| Outcome | Family | Registered contrast | N same-day (of record) | Late pairs excluded (delay, days) | Median change [95% CI] | Mean change | $d_z$ (band) | Hedges' g [95% CI] | W | p | Of record: median change, $d_z$, g, p |
|---|---|---|---|---|---|---|---|---|---|---|---|
| PSS-10 | F9 | intake to Day 5 | 10 (14) | 4 (1 to 4) | −11.0 [−17.0, −3.0] BCa | −10.80 | −1.56 (large) | −1.43 [−2.32, −0.86] BCa | 0 | 0.0020 | −12.0, −1.64, −1.54, 0.00096 |
| PANAS positive affect | F9 | intake to Day 5 | 10 (14) | 4 (1 to 4) | +10.0 [4.0, 15.5] BCa | +9.80 | 1.59 (large) | 1.45 [1.01, 2.30] BCa | 0 | 0.0020 | +13.0, 1.49, 1.40, 0.00098 |

| Outcome | Family | Registered contrast | N same-day (of record) | Late pairs excluded (delay, days) | Median change [95% CI] | Mean change | $d_z$ (band) | Hedges' g [95% CI] | W | p | Of record: median change, $d_z$, g, p |
|---|---|---|---|---|---|---|---|---|---|---|---|
| PANAS negative affect | F9 | intake to Day 5 | 10 (14) | 4 (1 to 4) | −14.0 [−18.0, −11.0] BCa | −15.50 | −1.71 (large) | −1.57 [−2.76, −0.89] BCa | 0 | 0.0020 | −12.5, −1.53, −1.44, 0.0012 |
| NGSE-8 | none | intake to Day 5 | 10 (14) | 4 (1 to 4) | +9.5 [5.0, 13.0] BCa | +9.80 | 1.55 (large) | 1.42 [0.77, 2.21] BCa | 0 | 0.0039 | +9.5, 1.64, 1.54, 0.0015 |
| AFTT specificity | F10 | intake to Day 5 | 4 (6) | 2 (1 to 2) | +0.079 [−0.090, 0.211] percentile | +0.0693 | 0.56 (moderate) | 0.41 [n.c.: degenerate bootstrap] percentile | 3 | 0.63 | +0.092, 0.88, 0.74, 0.16 |

**Table S4d.** The model of Table S4a fitted on the participants with an intake reading of the instrument, a sensitivity (Deviation 77), each timepoint contrast read on the t reference of Table S4b; the counts and the estimate on every participant in parentheses. The model assumes observations missing at random given the participant.

| Outcome | Participants | Observations | Term | Estimate | SE | df | 95% CI (t) | p (t) |
|---|---|---|---|---|---|---|---|---|
| CD-RISC-10 | 14 (18) | 55 (67) | +21 days against intake | +4.59 (+5.08) | 1.33 | 38 | [1.90, 7.28] t | 0.0014 |
| CD-RISC-10 | 14 (18) | 55 (67) | +14 days against intake | +4.07 (+4.59) | 1.30 | 38 | [1.45, 6.69] t | 0.0032 |
| CD-RISC-10 | 14 (18) | 55 (67) | Day 5 against intake | +8.36 (+7.42) | 1.30 | 38 | [5.74, 10.98] t | $1.4 \times 10^{-7}$ |
| PCL-5 | 15 (18) | 33 (37) | +21 days against intake | −3.40 (−4.13) | 3.20 | 16 | [−10.19, 3.39] t | 0.30 |
| PCL-5 | 15 (18) | 33 (37) | Early at-home post against intake | −9.19 (−9.90) | 5.37 | 16 | [−20.57, 2.20] t | 0.11 |
| PSS-10 | 14 (18) | 55 (67) | +21 days against intake | −4.25 (−4.88) | 1.46 | 38 | [−7.21, −1.30] t | 0.0059 |
| PSS-10 | 14 (18) | 55 (67) | +14 days against intake | −5.21 (−5.08) | 1.42 | 38 | [−8.10, −2.33] t | 0.00076 |
| PSS-10 | 14 (18) | 55 (67) | Day 5 against intake | −11.21 (−10.64) | 1.42 | 38 | [−14.10, −8.33] t | $1.6 \times 10^{-9}$ |
| PANAS positive affect | 14 (18) | 55 (67) | +21 days against intake | +3.78 (+3.81) | 1.60 | 38 | [0.54, 7.03] t | 0.024 |
| PANAS positive affect | 14 (18) | 55 (67) | +14 days against intake | +2.86 (+3.32) | 1.56 | 38 | [−0.31, 6.02] t | 0.076 |
| PANAS positive affect | 14 (18) | 55 (67) | Day 5 against intake | +11.21 (+10.87) | 1.56 | 38 | [8.05, 14.38] t | $1.5 \times 10^{-8}$ |
| PANAS negative affect | 14 (18) | 55 (67) | +21 days against intake | −5.86 (−7.12) | 1.97 | 38 | [−9.86, −1.86] t | 0.0052 |
| PANAS negative affect | 14 (18) | 55 (67) | +14 days against intake | −6.64 (−7.14) | 1.93 | 38 | [−10.55, −2.74] t | 0.0014 |
| PANAS negative affect | 14 (18) | 55 (67) | Day 5 against intake | −14.07 (−12.42) | 1.93 | 38 | [−17.97, −10.17] t | $9.7 \times 10^{-9}$ |
| WHO-5 (0 to 100 index) | 14 (18) | 55 (67) | +21 days against intake | +22.00 (+21.66) | 3.79 | 38 | [14.33, 29.68] t | $1.1 \times 10^{-6}$ |
| WHO-5 (0 to 100 index) | 14 (18) | 55 (67) | +14 days against intake | +18.57 (+20.56) | 3.70 | 38 | [11.08, 26.06] t | $1.2 \times 10^{-5}$ |
| WHO-5 (0 to 100 index) | 14 (18) | 55 (67) | Day 5 against intake | +37.43 (+35.67) | 3.70 | 38 | [29.94, 44.92] t | $2.4 \times 10^{-12}$ |
| NGSE-8 | 14 (18) | 41 (49) | +21 days against intake | +3.12 (+4.05) | 1.36 | 25 | [0.33, 5.92] t | 0.030 |
| NGSE-8 | 14 (18) | 41 (49) | Day 5 against intake | +9.43 (+8.71) | 1.32 | 25 | [6.70, 12.15] t | $1.8 \times 10^{-7}$ |

### S5. Descriptive figures

Fig. S2 shows the in-app daily check-ins and sleep quality across the retreat, without tests.

### S6. The retreat program

Table S6 gives the program of August 9 to 13, 2026, as the operator scheduled it, in general terms. Times are local.

**Table S6.** The program by day.

| Day | Time | Session |
|---|---|---|
| Day 1, August 9 | 18:00–19:00 | Arrival and settling in |
| | 19:00–20:30 | Dinner |

| Day | Time | Session |
| --- | --- | --- |
| | 20:30 | Opening group session and introductions |
| | 21:00–21:30 | Introduction to breathing practice |
| | 21:30–22:00 | Guided relaxation session |
| Day 2, August 10 | 08:00–09:30 | Breakfast |
| | 10:00–11:30 | Morning session: light movement and mindfulness practice |
| | 11:30–13:00 | Guided walk in the forest |
| | 13:00–13:30 | Guided meditation |
| | 14:00–16:00 | Lunch, then rest, journaling and personal time |
| | 16:00–17:00 | Guided nature session outdoors |
| | 17:00–19:00 | Sauna, optional |
| | 19:00–20:30 | Dinner |
| | 20:30 | Evening group gathering with breathing practice and a guided group reflection |
| | 20:45 | Guided relaxation session |
| | 21:30 | Closing group session for the day |
| Day 3, August 11 | 08:00–09:30 | Breakfast |
| | 10:00–11:00 | Morning movement and breathing practice |
| | 11:00–12:00 | Guided group reflection session in smaller groups, with breathing practice |
| | 12:00–13:30 | Lunch and preparation |
| | 13:30–15:00 | Cold-water immersion, optional |
| | 15:00–16:00 | Group session |
| | 16:00–18:00 | Sauna, rest and journaling |
| | 19:00–20:30 | Dinner |
| | 20:30 | Evening group gathering with breathing practice |
| | 22:00 | Closing group session for the day |
| Day 4, August 12 | 08:00–09:30 | Breakfast |
| | 10:00–10:45 | Morning movement and breathing practice |
| | 11:00–12:30 | Guided group reflection session |
| | 12:30–13:10 | Guided meditation |
| | 13:10–14:00 | Lunch |
| | 14:00–16:00 | Rest, journaling and personal time |
| | 16:00–17:15 | Movement practice outdoors in nature |
| | 17:15–19:00 | Sauna, optional, and quiet time |
| | 19:00 | Dinner |
| | 20:30 | Evening group gathering, with breathing practice |
| | 22:00 | Closing session for the day |
| Day 5, August 13 | 08:00–09:30 | Breakfast |
| | 10:00–11:00 | Morning movement and group sharing |
| | 11:00–12:30 | Closing group session |
| | | After the closing session, an outing to the city, then departure for the airport |

The sauna and the cold water were optional, and joining any session was each participant's own choice. The sessions were led by three facilitators: the program's founder, certified in a nature-based wellbeing method; a clinical psychologist; and an art therapist working through nature. Every session was held with the whole group, except the reflection session of Day 3, held in smaller groups, and the individual journaling time.

The app during the retreat, as it was scheduled: participants were asked to be phone-free during sessions; the app was used between them. Day 1: one brief check-in in the evening, at 22:00. Days 2 to 5: the sleep diary each morning, the first time the app was opened. Days 2 to 4: a brief check-in at 09:00, one at midday (Day 2 at 14:45, Days 3 and 4 at 12:30) and one in the evening (Day 2 at 22:30, Day 3 at 22:15, Day 4 at 22:30). Day 5: a brief check-in at 09:00, then the closing questionnaires at 09:30. Every check-in and questionnaire, but not the sleep diary, opened with a brief guided breathing exercise.

### Figure captions

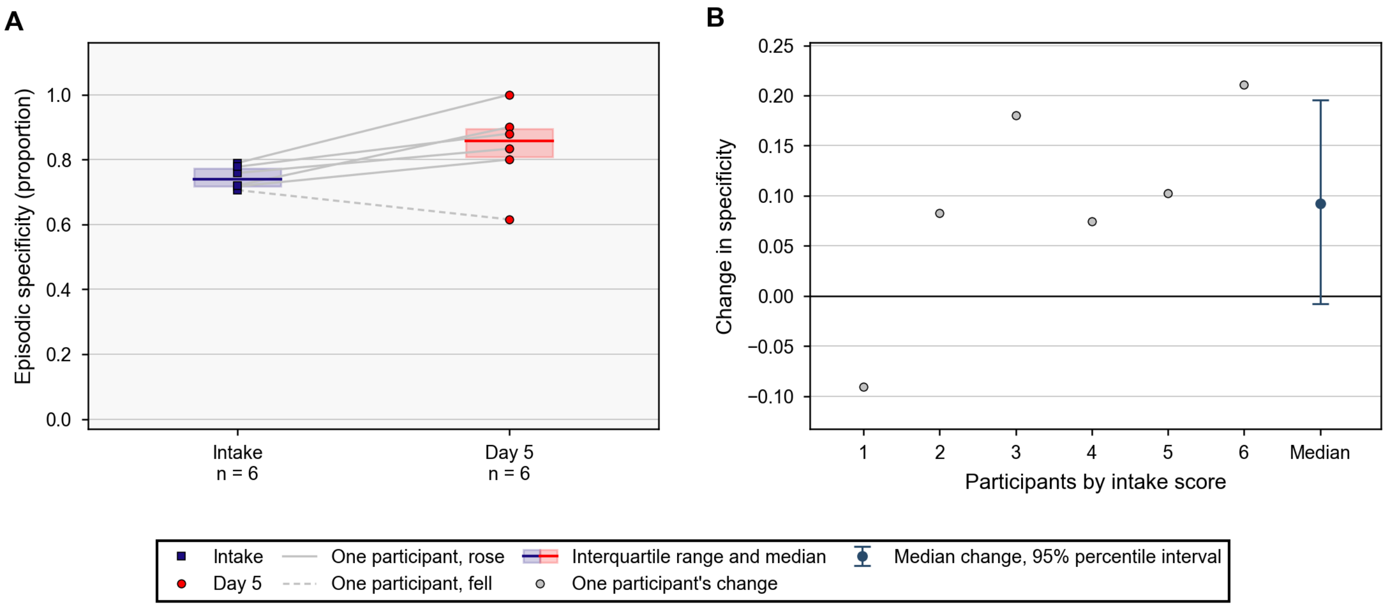


**Fig. S1.** Episodic specificity on the AFTT at intake and Day 5. The score is the share of coded details that are internal (specific to the imagined event). A, each participant's readings (N = 6). Intake squares and Day 5 circles are joined by a gray line, solid where the score rose (5) and dashed where it fell (1). The interquartile range is shaded and its median is a line. B, the changes by intake score, with their median and its 95% percentile interval and a solid zero line. The median change in specificity from intake to Day 5 was +0.092 (95% percentile CI −0.008 to 0.195); Hedges' g = 0.74 (95% percentile CI 0.09 to 3.04); Wilcoxon W = 3, p = 0.16. Abbreviations: AFTT, autobiographical future-thinking task.

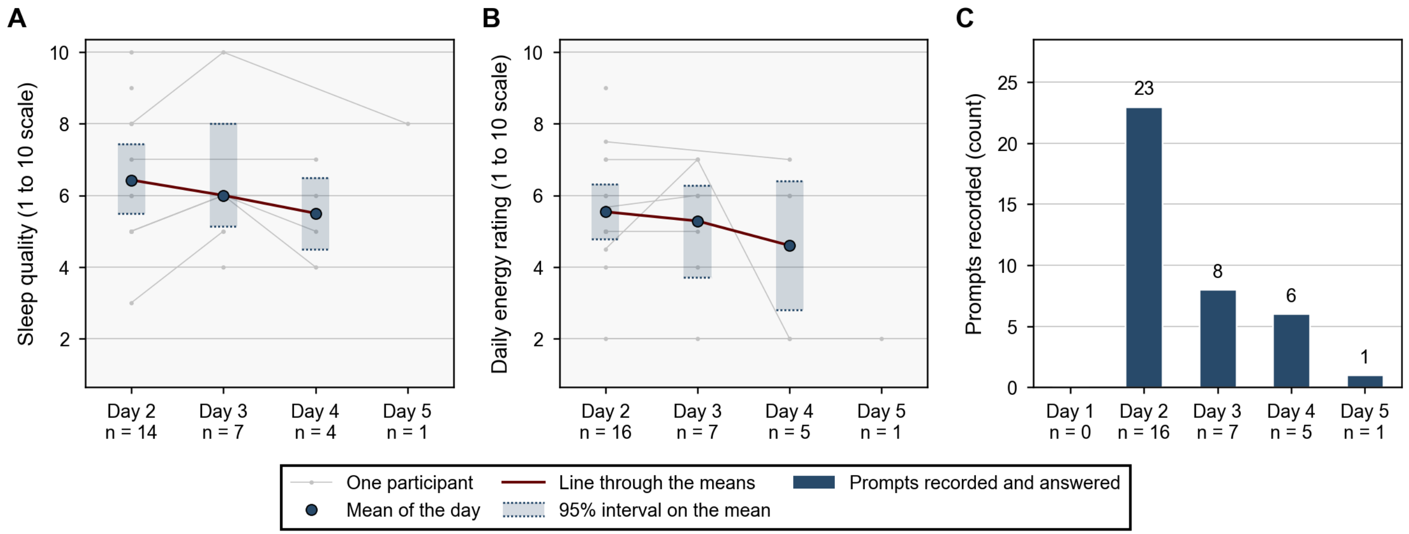


**Fig. S2.** The in-app daily check-ins across the retreat. A, sleep quality, and B, the daily energy rating, each on its 1 to 10 scale. Gray lines join each participant's ratings. The circles are the means of the day, drawn where at least 3 participants rated, each with its 95% interval on the mean shaded between dotted bounds (BCa bootstrap; percentile where a day's N is below 7). A line joins the means. C, the number of prompts recorded each day; every prompt was answered. Panels A and B span Days 2 to 5, neither item being scheduled on Day 1; panel C, the prompts recorded, spans Days 1 to 5. Day labels give the participants who rated that item that day (in C, those with a record); 16 of the N = 18 study participants recorded 29 participant-days. No test is shown. Abbreviations: BCa, bias-corrected and accelerated.
